# Genetic variation near CXCL12 is associated with susceptibility to HIV-related non-Hodgkin lymphoma

**DOI:** 10.1101/19011999

**Authors:** Christian W. Thorball, Tiphaine Oudot-Mellakh, Christian Hammer, Federico A. Santoni, Jonathan Niay, Dominique Costagliola, Cécile Goujard, Laurence Meyer, Sophia S. Wang, Shehnaz K. Hussain, Ioannis Theodorou, Matthias Cavassini, Andri Rauch, Manuel Battegay, Matthias Hoffmann, Patrick Schmid, Enos Bernasconi, Huldrych F. Günthard, Paul J. McLaren, Charles S. Rabkin, Caroline Besson, Jacques Fellay

## Abstract

Human immunodeficiency virus (HIV) infection is associated with a substantially increased risk of non-Hodgkin lymphoma (NHL). High plasma viral load, low CD4+ T cell counts and absence of antiretroviral treatment (ART) are known predictive factors for NHL. Even in the era of suppressive ART, HIV-infected individuals remain at increased risk of developing NHL compared to the general population. To search for human genetic determinants of HIV-associated NHL, we performed case-control genome-wide association studies (GWAS) in three cohorts of HIV+ patients of European ancestry and meta-analyzed the results. In total, 278 cases and 1924 matched controls were included. We observed a significant association with NHL susceptibility in the C-X-C motif chemokine ligand 12 (*CXCL12*) region on chromosome 10. A fine mapping analysis identified rs7919208 as the most likely causal variant (P = 4.77e-11). The G>A polymorphism creates a new transcription factor binding site for BATF and JUND. Analyses of topologically associating domains and promoter capture Hi-C data revealed significant interactions between the rs7919208 region and the promoter of *CXCL12*, also known as stromal-derived factor 1 (*SDF-1*). These results suggest a modulatory role of *CXCL12* regulation in the increased susceptibility to NHL observed in the HIV-infected population.

## Introduction

Human immunodeficiency virus (HIV) infection is associated with a markedly increased risk of several types of cancer compared to the general population.^1–3^ This elevated cancer risk can be attributed partly to viral-induced immunodeficiency, frequent co-infections with oncogenic viruses (e.g., Epstein-Barr virus (EBV), hepatitis B and hepatitis C viruses, human herpesvirus 8 (HHV-8) and papillomavirus), and increased prevalence of traditional risk factors such as smoking.^4,5^ However, all of these risk factors may not entirely explain the excess cancer burden seen in the HIV+ population.^6^

A previous study performed in the Swiss HIV Cohort Study (SHCS) identified two AIDS-defining cancers, Kaposi sarcoma and non-Hodgkin lymphoma (NHL) as the main types of cancer found among HIV positive patients (NHL representing 34% of all identified cancers).^4^ The relative risk of developing NHL in HIV patients was highly elevated compared to the general population (period-standardized incidence ratio (SIR) = 76.4).^4^ High HIV plasma viral load, absence of antiretroviral therapy (ART) as well as low CD4+ T cell counts are known predictive factors for NHL.^7,8^ The introduction of ART into clinical practice has led to improved overall survival and restoration of immunity by decreasing viral load and increasing CD4+ T cell counts, and has led to a decreased risk of developing NHL. However, the risk remains substantially elevated compared to the general population (SIR = 9.1 (8.3–10.1))^9^ and NHL still represents 20% of all cancers in people living with HIV in the ART era.^10^ Non-Hodgkin’s lymphomas associated with HIV are predominantly aggressive B-cell lymphomas. Although they are heterogeneous, they share several pathogenic mechanisms involving chronic antigen stimulation, impaired immune response, cytokine deregulation and reactivation of the oncogenic viruses EBV and HHV-8^11^

The emergence of genome-wide approaches in human genomics has led to the discovery of many associations between common genetic polymorphisms and susceptibility to several diseases including HIV infection and multiple types of cancer.^12,13^ Recent genome-wide association studies (GWAS) of NHL have identified multiple susceptibility loci in the European population.^14–22^ These variants are located in the genes *LPXN*^21^, *BTNL2*^23^, *EXOC2, NCOA1*^14^, *PVT1*^14,22^, *CXCR5, ETS1, LPP*, and *BCL2*^22^ for various subtypes of NHL, as well as *BCL6* in the Chinese population.^24^ Strong associations with variation in human leukocyte antigen (HLA) genes have also been reported.^15,18,22^ However, in the setting of HIV infection, no genome-wide analysis has been reported concerning the occurrence of NHL and the specific mechanisms driving their development remain largely unknown.

Here we report the results of the first genome-wide analysis of NHL susceptibility in individuals chronically infected with HIV. We combined three HIV cohort studies from France, Switzerland and the USA and searched for associations between >6 million single nucleotide polymorphisms (SNPs) and a diagnosis of NHL. We identified a novel genetic locus near *CXCL12* as associated with the development of NHL among HIV+ individuals.

## Materials and methods

### Ethics statement

The Swiss HIV Cohort Study (SHCS), the Primo ANRS and ANRS CO16 Lymphovir cohorts (ANRS) and the Multicenter AIDS Cohort Study (MACS) cohorts have been approved by the competent ethics committees / institutional review boards of all participating institutions. A written informed consent, including consent for human genetic testing, was obtained from all study participants.

### Study participants and contributing centers

#### Swiss HIV Cohort Study (SHCS)

The SHCS is a large, ongoing, multicenter cohort study of HIV-positive individuals that includes >70% of adult living with HIV in Switzerland. At follow-up visits every 6 months, demographic, clinical, laboratory, and ART information has been prospectively recorded since 1988.^25^ Cancer diagnoses are verified thoroughly using checking charts including information on biopsies and imaging. To minimize potential treatment bias and population stratification, we only considered as cases patients diagnosed with NHL between 2000 and 2017 and of European ancestry, as determined by principal component analysis (PCA) (supplemental Figure 1A). Controls were matched based on age, ancestry, CD4+ T cell counts and viral load results. To be eligible as controls, they also had to be diagnosed with HIV prior to 2005 and have no registered cancer diagnosis of any type as of 2017. Patients were genotyped using Illumina HumanOmniExpress-24 Beadchips, or genotypes were obtained in the context of a previous GWAS in the SHCS on various platforms including Illumina HumanCore-12, HumanHap550, Human610 and Human1M Beadchips.

#### French Primo ANRS and ANRS CO16 Lymphovir cohorts (ANRS)

The French ANRS CO16 lymphovir cohort of HIV related lymphomas enrolled adult patients at diagnosis of lymphoma in 32 centers between 2008 and 2015.^26^ Pathological materials were centralized, and diagnoses of NHL were based on World Health Organization criteria. Patients were genotyped using Illumina Human Omni5 Exome 4v beadchips. Additional cases and controls were included from the ANRS PRIMO Cohort, which has been enrolling patients during primary HIV-1 infection in 95 French Hospitals since 1996.^27^ Patients were genotyped using Illumina Sentrix Human Hap300 Beadchips. Only patients of European ancestry, as determined by PCA, were included in the study (supplemental Figure 1B).

#### The Multicenter AIDS Cohort Study (MACS)

The MACS has enrolled gay and bisexual HIV infected men in 4 US cities since 1984. The NHL cases were predominately diagnosed prior to the year 2000. Data collected include demographic variables (age, race, ethnicity and HIV transmission category), CD4+ T cell count, HIV viral load and tumor histology. Eligible cases had a diagnosis of HIV-related NHL, available genotyping data and at least one CD4+ T cell count obtained within 2 years of the NHL diagnosis. Controls were matched on MACS study site, age at NHL diagnosis (+/- 2 years) and CD4+ T cell count at NHL diagnosis (within the following groups 0-99 / 100 -199 / 200-499 / >499 cells/µL). Patients were genotyped using Illumina HumanHap550 and Human1M Beadchips.^28^ As in the other cohorts, only individuals of European ancestry were included, as determined by PCA (supplemental Figure 1C).

### Quality control and imputation of genotyping data

The genotyping data from each cohort was filtered and imputed in a similar way, with each genotyping array processed separately to minimize potential batch effects. All variants were first flipped to the correct strand orientation with BCFTOOLS (v1.8) using the human genome build GRCh37 as reference. Variants were removed if they had a larger than 20% minor allele frequency (MAF) deviation from the 1000 genomes phase 3 EUR reference panel or if they showed a larger than 10% MAF deviation between genotyping chips in the same cohort.

The QC filtered genotypes were phased with EAGLE2^29^ and missing genotypes were imputed using PBWT^30^ with the Sanger Imputation Service^31^, taking the 1000 Genomes Project Phase 3 panel as reference. Only high-quality variants with an imputation score (INFO > 0.8) were retained for further analyses.

### Genome-wide association testing and meta-analysis

To search for associations between human genomic variation and the development of HIV-related NHL, we first performed separate GWAS within each cohort (SHCS, ANRS and MACS) prior to combining the results in a meta-analysis.

For each cohort separately, the imputed variants were filtered out using PLINK (v2.00a2LM)^32^ based on missingness (> 0.1), minor allele frequency (< 0.02) and deviation from Hardy-Weinberg Equilibrium (PHWE < 1e-6). Determination of population structure and calculation of principal components was done using EIGENSTRAT (v6.1.4)^33^ and the HapMap3 reference panel^34^. All individuals not clustering with the European HapMap3 samples were excluded from further analyses. The samples were screened using KING (v2.1.3)^35^ to ensure no duplicate or cryptic related samples were included. Single-marker case-control association analyses were performed using linear mixed models, with genetic relationship matrices calculated between pairs of individuals according to the leave-one-chromosome-out principle, as implemented in GCTA mlma-loco (v1.91.4beta).^36,37^ Sex was included as a covariate, except in the MACS cohort, which only includes men.

The results of the three GWAS were combined across cohorts using a weighted Z-score-based meta-analysis in PLINK (v1.90b5.4), after exclusion of the variants that were not present in all three cohorts.

### Fine mapping of associated regions

Fine mapping of the *CXCL12* locus was performed using PAINTOR (v3.1)^38^ to identify the most likely causal variant(s). All variants within 200kb of the top associated SNP and with a p-value below 0.005 were included in the model. The linkage disequilibrium (LD) matrix was created using PLINK and genotype data from the SHCS cohort. PAINTOR was first run against all genomic annotation databases provided with the software, including the FANTOM5, ENCODE and the Roadmap Epigenomics Project. For the final model, the top 5 annotations based on improvement to model fit and cell type relevance were selected to obtain the posterior probabilities and the 99% credible set of the variants most likely to be causal based on the association from Bayes’ factors.

### Predictive effect of potentially causal variants

The potential functional impact of the predicted causal variants was assessed using DeepSEA^39^, a deep learning-based sequence model trained on available chromatin and transcription factor data from ENCODE and Roadmap Epigenomics. DeepSEA provides a functional significance score for each variant, which is a measure of the evolutionary conservation and the significance of the magnitude of the predicted chromatin effects. For the variants with a functional significance score of less than 0.01, we analyzed the predicted changes in specific chromatin modifications or transcription factor (TF) binding probabilities. Chromatin or TF binding changes with E-values below 0.001 and normalized probabilities of observing a binding event above 0.2 were considered relevant. The TF position weight matrices (PWMs) for TFs with a high probability of binding (normalized probability ≥ 50%) were obtained from the JASPAR CORE 5.0 database.^40^

### Long-range chromatin interactions

Predicted topological associating domains (TADs) near the genome-wide significant locus in GM12878 lymphoblastoid cells were obtained from publicly available data^41^ and visualized using the 3D Genome Browser.^42^

Potential interactions between the genome-wide significant locus and promoters of nearby genes were analyzed using publicly available promoter capture Hi-C data in GM12878 lymphoblastoid cells. The Hi-C data was processed through the CHiCAGO pipeline and visualized with CHiCP.^43,44^ Interaction scores ≥ 5 were considered significant, as described previously.^45^

### Expression quantitative trait loci (eQTL) analyses

The role of rs7919208 as an eQTL was examined in GEUVADIS^46^ and in response to various pathogens, although not including HIV, in the Milieu Intérieur Consortium cohort.^47^ Furthermore, eQTL information was also obtained from the GTEx (v7)^48^ Portal on 03/22/2019.

Bulk RNA Barcoding and sequencing (BRB-seq)^49^ was performed on RNA from peripheral blood mononuclear cells (PBMCs) of 452 individuals from the SHCS with available genotyping data.

### Comparison to GWAS hits in the general population

An attempt at replicating variants previously associated with NHL in the general population was performed by extraction of the p-values of the SNPs reported to be associated in previous NHL GWAS. A variant was considered replicated if it had a nominally significant association p-value (P < 0.05) plus similar effect direction in the meta-analysis.

The effect of rs7919208 in the general population cohorts was assessed directly using the NIH database for Genotypes and Phenotypes (dbGaP) accession # phs000801 cohorts for chronic lymphocytic leukemia (CLL), DLBCL (Diffuse large B-cell lymphoma), FL (Follicular lymphoma) and MZL (Marginal zone lymphoma) and corresponding controls.^14,22,23,50^ The genotype data was imputed, processed and analyzed using the same pipeline and methods as described above for the HIV cohorts, with duplicate samples identified and removed using KING and including age and sex as covariates.

### Statistical analyses

All statistical analyses were performed using the R statistical software (v3.3.3), unless otherwise specified.

### Data sharing statement

Full summary statistics will be made available in the GWAS catalog (https://www.ebi.ac.uk/gwas) upon publication. The raw genotype data can be obtained through the respective cohorts.

## Results

### Study participants and association testing

To identify human genetic determinants of HIV-associated NHL, we performed case-control genome-wide association studies (GWAS) in three groups of HIV+ patients of European ancestry (SHCS, ANRS and MACS). The characteristics of the study participants are presented in Table 1. In total, genotyping data were obtained for 278 cases (NHL+/HIV+) and 1924 matched controls (NHL-/HIV+). With this sample size, we had 80% power to detect a common genetic variant (10% minor allele frequency) with a relative risk of 2.5, assuming an additive genetic model and using Bonferroni correction for multiple testing (P_threshold_ = 5e-8).^51^

**Table 1.**
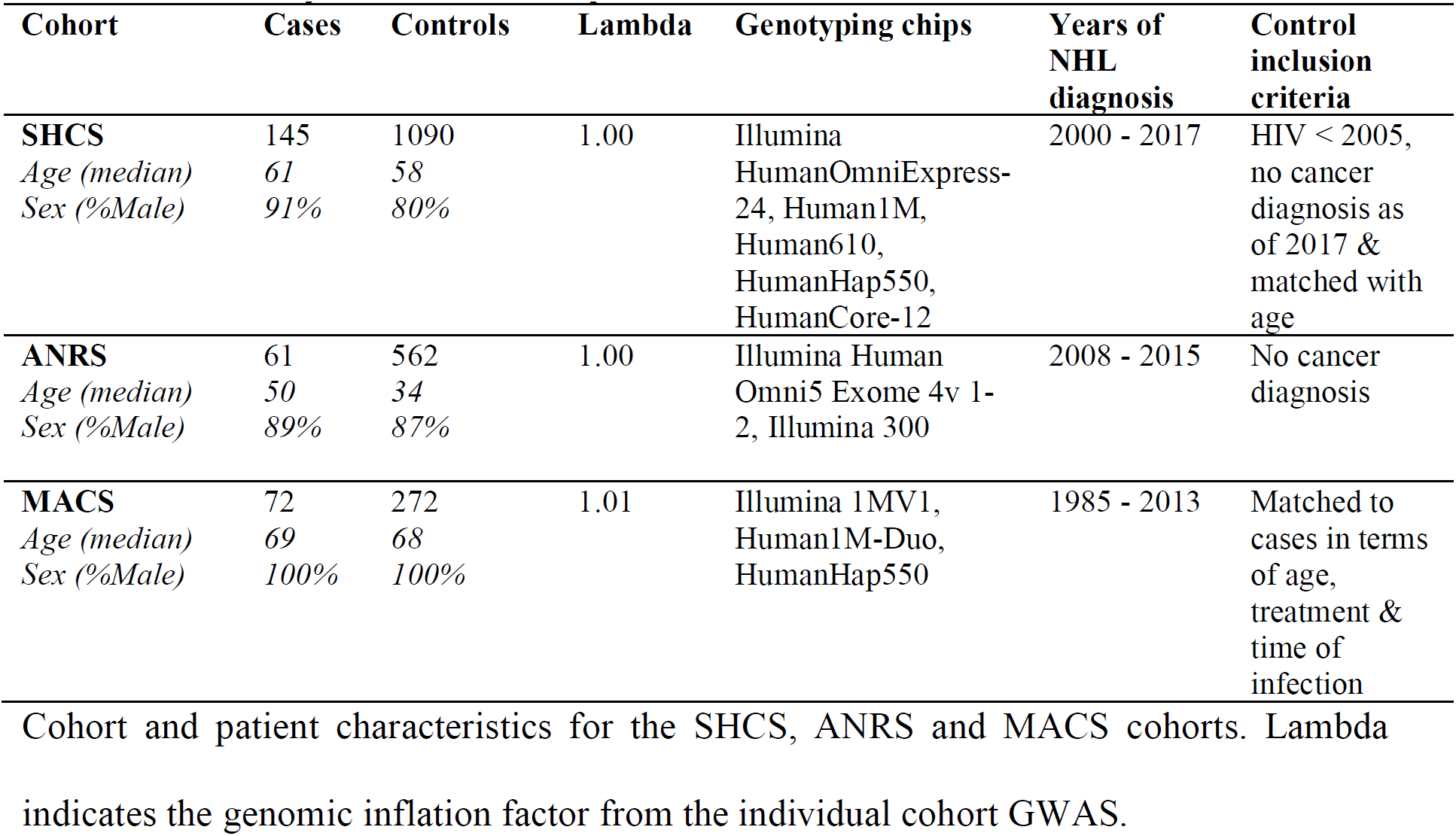
Summary of included samples and studies.

After genome-wide imputation and quality control, 6.2 million common variants were tested for association with the development of NHL using linear mixed models including sex as a covariate. Results were combined across cohorts using a weighted Z-score-based meta-analysis (Figure 1A). The genomic inflation factor (lambda) was in all cases very close to 1 [1.00–1.01], indicating an absence of systematic inflation of the association results (Figure 1B; supplemental Figure 2).

**Figure 1.**
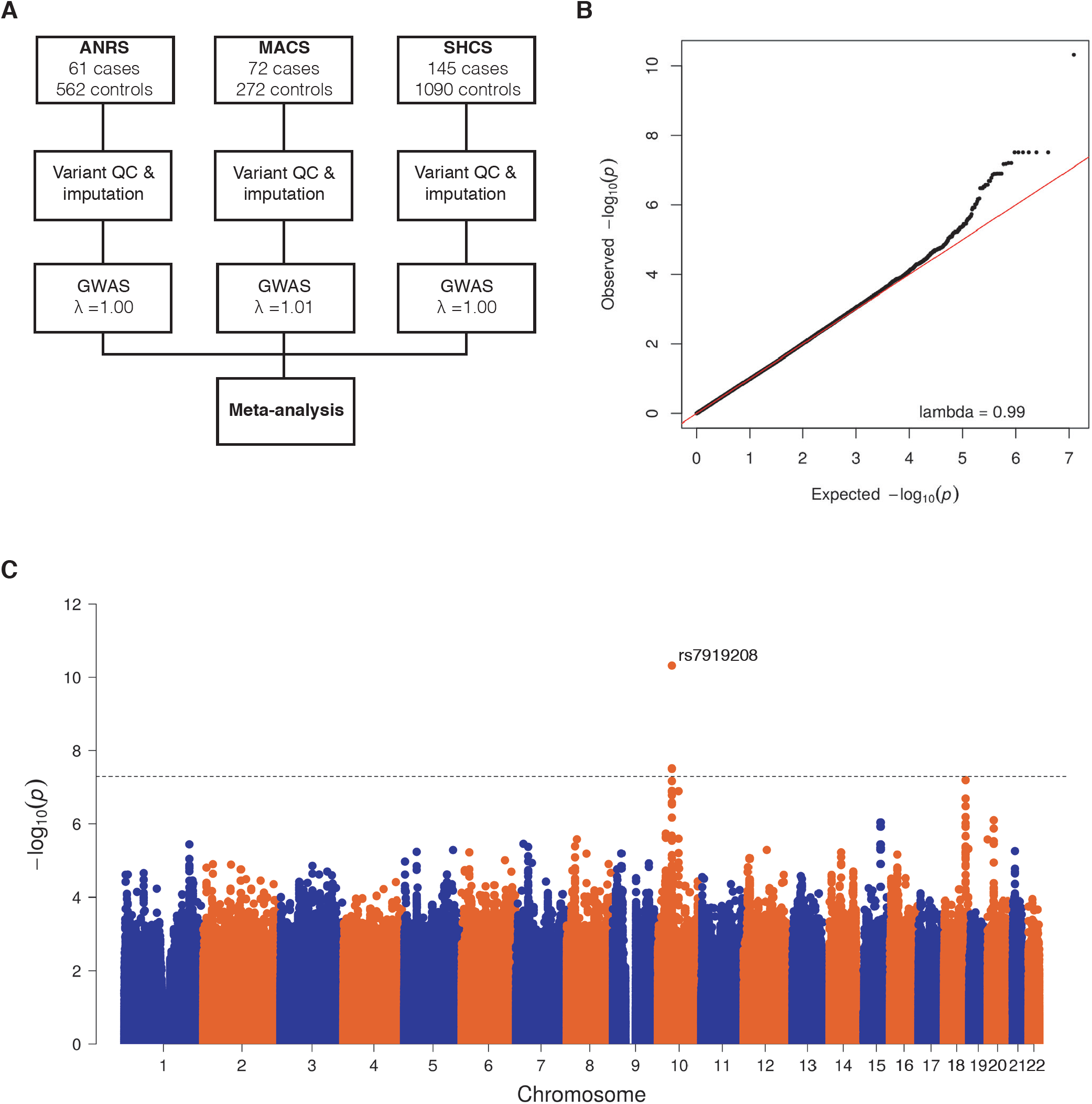
Genome-wide association analysis. (A) Schematic of analysis pipeline. (B) Quantile-quantile plot of the observed -log10(p-value) (black dots, y-axis) versus expected -log10(p-values) under the null hypothesis (red line) to check for any genomic inflation of the observed p-values. No genomic inflation is observed, with the genomic inflation factor lambda = 0.99. (C) Manhattan plot of all obtained p-values for each variant included in the meta-analysis. The genome-wide threshold (P = 5e-8) for significance is marked by a dotted line. Only variants at the CXCL12 locus were found to be significant.

### Association results

We observed significant associations with the development of HIV-related NHL at a single locus on chromosome 10, downstream of *CXCL12* (Figure 1C). A total of 7 SNPs in this locus had p-values lower than the genome-wide significance threshold (P < 5e-8), with rs7919208 displaying the strongest association (Table 2). This association was only detected in the SHCS and ANRS cohorts and not among MACS study participants (supplemental Table 1).

**Table 2.** Significant association with HIV-related NHL.

| Chr | Pos | SNP | Ref | Alt | P | OR |
| --- | --- | --- | --- | --- | --- | --- |
| 10 | 44673557 | rs7919208 | A | G | 4.77e-11 | 1.23 |
| 10 | 44677967 | rs149399290 | T | C | 3.09e-08 | 1.20 |
| 10 | 44678218 | rs17155463 | T | A | 3.09e-08 | 1.20 |
| 10 | 44678262 | rs17155474 | C | T | 3.09e-08 | 1.20 |
| 10 | 44678454 | rs17155478 | T | C | 3.09e-08 | 1.20 |
| 10 | 44678898 | rs12249837 | G | A | 3.09e-08 | 1.20 |
| 10 | 44680902 | rs10608969 | T | TAAAGA | 3.09e-08 | 1.20 |

Variants significantly associated with HIV-related NHL in a weighted Z-score-based meta-analysis of all individuals included in the SHCS, ANRS and MACS cohorts. Odds ratios (OR) were transformed from betas using the formula $OR = \exp(\text{beta})$ .

### Fine mapping of the CXCL12 locus

To identify the causal variant(s) among associated SNPs and determine their potential functional effects, we used a multi-level fine mapping approach, combining the statistical fine mapping tool PAINTOR to obtain a 99% credible set and the deep learning framework DeepSEA to predict any effects on chromatin marks and transcription factor binding these variants may have.

Using PAINTOR, we identified a single variant, rs7919208, having a high posterior probability (= 100%) of being causal among the 99% credible set based on the integration of the association results, LD structure and enrichment of genomic features in this locus (Figure 2).

**Figure 2.**
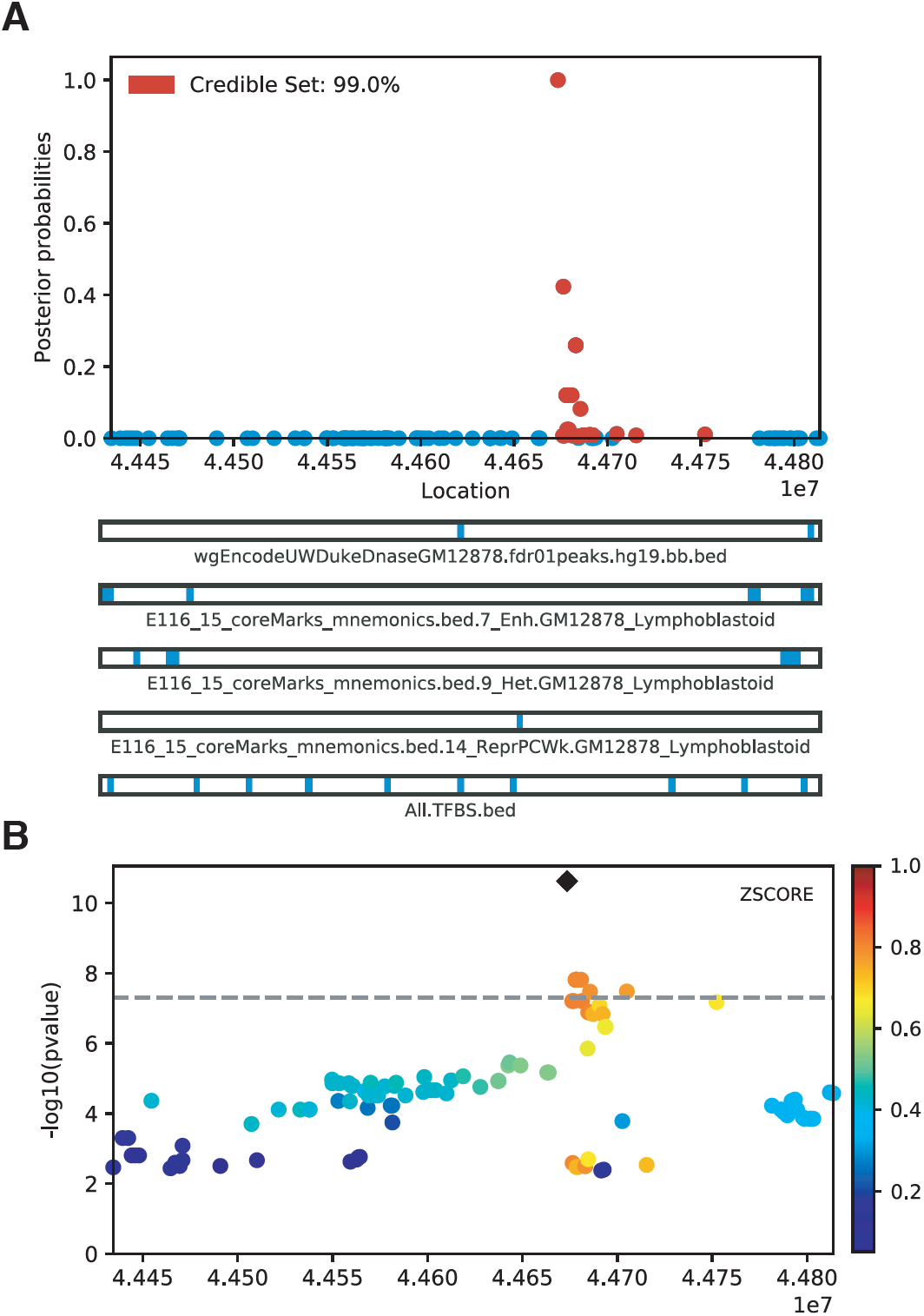
Fine mapping of genome-wide significant hits with PAINTOR. (A) The 99% credible set and posterior probabilities of being the causal variant. The genomic positions are listed on the x-axis. Bottom tracks represent DNAase and chromatin marks obtained from GM12878 cells as well as TFBS from the Roadmap Epigenomics Project and ENCODE in the region. (B) Locus plot of the associated variants, highlighting the LD relationship, based on the SHCS cohort. The top variant rs7919208 is marked by a black diamond.

Consistent with the PAINTOR result, DeepSEA also identified rs7919208 as the sole variant, among the 99% credible set, predicted to have a functional impact by significantly increasing the probability of binding by the B cell transcription factors BATF (log2 fold-change = 3.27) and JUND (log2 fold-change = 2.91) (supplemental Table 2). Further analysis of the genomic sequence surrounding rs7919208 and the JASPAR transcription factor binding site (TFBS) motifs for BATF and JUND revealed that rs7919208 G->A polymorphism creates the TFBS motif required for the binding of these transcription factors (Figure 3A).

**Figure 3.**
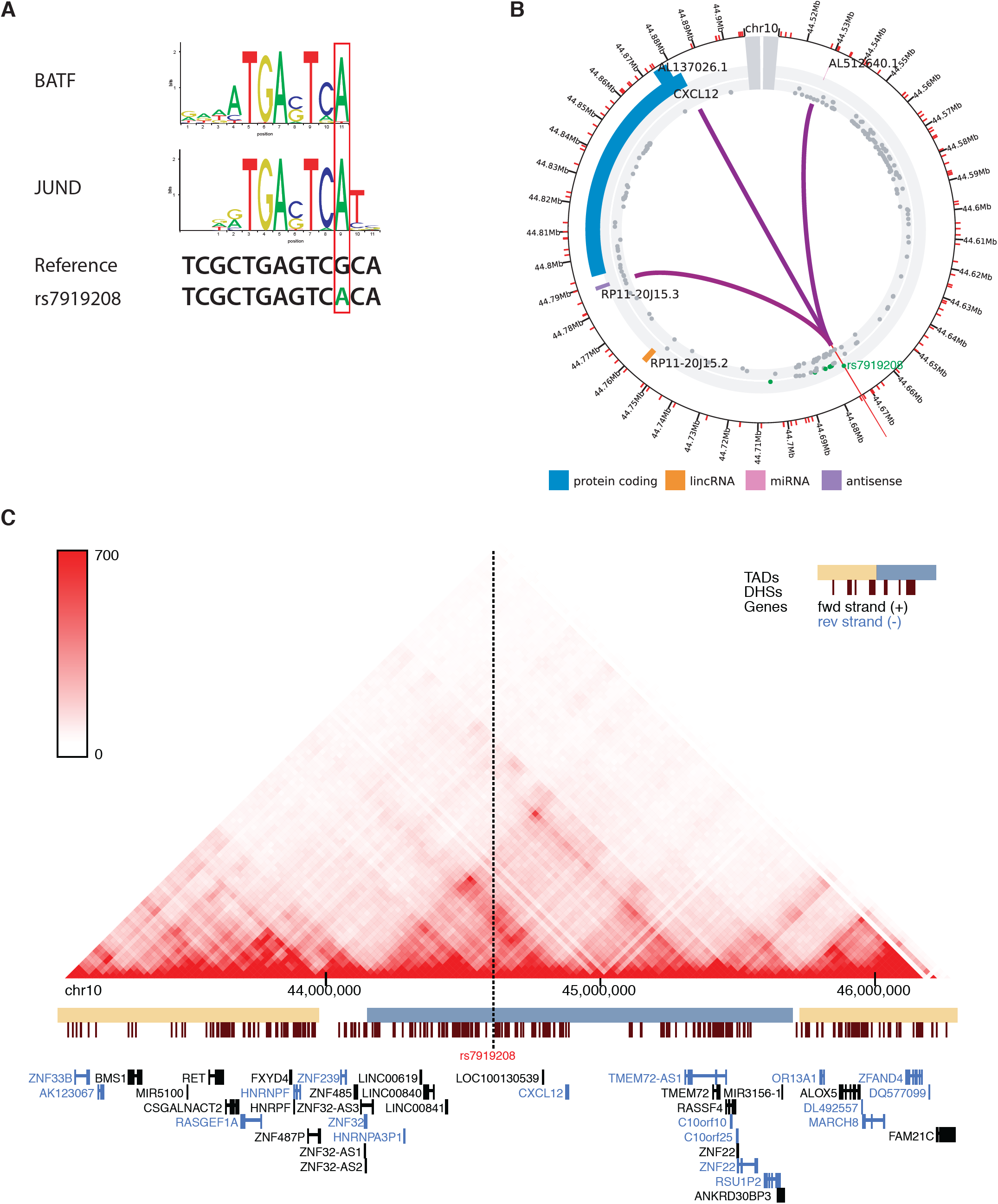
Novel transcription factor binding site and long-range interactions. (A) Canonical motifs of BATF and JUND with the underlying genomic reference sequence and the nucleotide change caused by rs7919208. (B) Promoter capture Hi-C analysis in the GM12878 cell line of the region with the predicted causal variant and CXCL12. Variants and their level of association in the meta-analysis are marked in the inner grey circle. Genome-wide significant variants are colored green. Purple lines indicate significant interactions between promoter and other genomic regions. (C) TADs in the GM12878 cell line in the region of CXCL12. The yellow and blue boxes indicate the called TADs from the Hi-C contact map above. The plot is centered on rs7919208.

### Long-range chromatin interactions

To assess the potential functional links between the TFBS created in the presence of the minor allele of rs7919208 and the nearby genes, we performed an analysis of promoter capture Hi-C data and topologically associating domains (TADs). We used the well-characterized GM12878 lymphoblastoid cell line produced by EBV transformation of B lymphocytes collected from a female European donor as model.

First, to examine the interaction potential of the rs7919208 region with nearby promoters, we analyzed available promoter capture Hi-C data obtained from the GM12878 cell line. This analysis revealed a significant interaction between the rs7919208 region and the *CXCL12* promoter, suggesting a possible modulating impact of rs7919208 on the transcription of that gene (Figure 3B). Second, to further validate this observed genomic interaction, we analyzed available TAD calls from GM12878 cells ^41^, using the 3D Genome Browser for visualization^42^ (Figure 3C). We observed that rs7919208 is located within a large TAD together with *CXCL12*, signifying the interaction potential of the new TFBS at rs7919208 and *CXCL12*.

### Transcriptomic effects of rs7919208

We did not observe any association between rs7919208 and mRNA expression levels of *CXCL12* in peripheral blood or PBMCs from multiple publicly available datasets, including GTEx (v7)^48^, GEUVADIS ^46^ and the Milieu Intérieur Consortium^47^ (supplemental Figure 3). Of note, *CXCL12* expression levels were very low in all datasets.

HIV infection causes many profound transcriptomic changes.^52^ Thus, in order to examine the effect of rs7919208 on *CXCL12* in the context of HIV infection, we extracted RNA from PBMCs of 452 individuals in the SHCS with available genotyping data and sequenced them using the Bulk RNA Barcoding and sequencing (BRB-seq) approach.^49^ However, the expression levels of *CXCL12* were below the limit of detection for most individuals, preventing an eQTL analysis.

### No replication of susceptibility loci found in the general population

To assess whether the genetic contribution to the risk of developing NHL is similar or distinct in the HIV+ population compared to the general population, we extracted the p-values of all variants found to be genome-wide significant in previous GWAS performed in the general population^14,21–24,53^ and compared them to our results. We did not replicate any of the previously published genome-wide associated variants, even at nominal significance level (P < 0.05), despite sufficient statistical power for many of the variants, thus indicating that the genetic susceptibility of NHL is distinct between the HIV+ and the general population (supplemental Table 3). To further examine this possibility, we tested whether the NHL/HIV+ associated variant rs7919208 is associated with an increased risk of NHL in the general population. We performed a series of case/control GWAS of four NHL subtypes (CLL, DLBCL, FL and MZL) as well as a combined GWAS with all NHL subtypes (supplemental Table 4; supplemental Figure 4) and assessed the association evidence at rs7919208. We found no association between rs7919208 and any of the subtypes in the general population, even at nominal significance.

## Discussion

In this genome-wide analysis, including a total of 278 NHL HIV+ cases and 1924 HIV+ controls from three independent cohorts, we identified a novel NHL susceptibility locus on chromosome 10 near the *CXCL12* gene. The strong signal observed in the meta-analysis was driven by the associations detected in the SHCS and ANRS cohorts and there was no evidence of association in the MACS cohort. Notably, most NHL cases in the MACS cohort date back to the pre-ART era, while only NHL cases diagnosed after the year 2000 were included in the SHCS and ANRS analyses. Conceivably, NHL occurring in the early years of the HIV pandemic may have been primarily driven by severe immunosuppression, which could have obscured any influence of human genetic variation among the cases in the MACS sample. Precise phenotype definition is crucial in designing large-scale genetic studies since any environmental noise tends to decrease the likelihood of identifying potential genetic influences.

NHL is a relatively rare cancer even among HIV infected individuals, making it difficult to collect the large numbers of cases that would typically be included in contemporary genome-wide genetic studies. Indeed, a recent study from the Data Collection on Adverse events of Anti-HIV Drugs (D:A:D) group showed an NHL incidence rate of 1.17/1000 person-years of follow-up over the past 15 years (392 new cases in >40,000 HIV-infected individuals).^8^ Still, we were able to obtain clinical and genetic data from a total of 278 patients with confirmed NHL diagnosis. By matching them with a larger number of controls from the same cohorts, we had enough power to identify associated variants of relatively large effects in the *CXCL12* region.

Several groups have already suggested a potential role for *CXCL12* variation in HIV-related NHLs. A prospective study correlated increased *CXCL12* expression with subsequent NHL development in HIV-infected children but not in uninfected children.^54^ The number of A alleles at the CXCL12-3’ variant (rs1801157) has also previously been associated with an increased risk of developing HIV-related NHL during an 11.7 year follow-up period.^55^ Thus, our data further support the role of CXCL12 as a critical modulator of the individual risk of developing NHL in the HIV population.

The role of CXCL12 and its receptor chemokine receptor 4 (CXCR4) in cancer in the general population is well established, with the levels of *CXCL12* and *CXCR4* found to be increased in multiple types of cancer and to be associated with tumor progression.^56,57^ Furthermore, *in vivo* inhibition of either CXCR4 or CXCL12 signaling is capable of disrupting early lymphoma development in severe combined immunodeficient (SCID) mice transfused with EBV+ PBMCs.^58^ These results and others have already led to the development and testing of several small molecules targeting either CXCL12 or CXCR4 to inhibit tumor progression.^56^

We could not identify any significant relationship between rs7919208 and the expression levels of *CXCL12* in PBMCs or EBV transformed lymphocytes. This can be due to multiple factors such as the low expression levels of *CXCL12* in most tissues, aside from stromal cells, or that rs7919208 through creation of the BATF and JUND binding site represent an induced or dynamic eQTL. These types of eQTLs are often found in regions deprived of regulatory annotations, since these have been examined in static cell types.^59^ HIV-induced overexpression of *BATF*^60^ could also explain why rs7919208 is only a risk factor in the HIV population and not in the general population.

Previous analyses in the general population have discovered both shared and distinct associations for NHL subtypes.^14,21–24,53^ However, similar analyses were not possible in our sample since NHL subtype information was not available for many of our cases. Furthermore, information on serostatus for relevant co-infections with EBV or other oncogenic viruses was not available and could therefore not be assessed. In particular, EBV has been largely associated with the development of NHL and other lymphomas and is considered a driver of a subset of NHLs in the general population.^61^ Variants in the HLA region have consistently been associated with all NHL subtypes in HIV uninfected populations regardless of EBV serostatus. We did not find any evidence of HLA associations in our analyses of HIV-related NHL. This lack of replication of HLA variants and of all other previously identified risk variants from the general population suggests that distinct genes or pathways influence susceptibility to NHL in the HIV+ population compared to the general population.^62^

In summary, we have identified variants significantly associated with the development of NHL in the HIV population. Fine mapping of the associated locus and subsequent analyses of TADs, promoter capture Hi-C data as well as deep-learning models of mutational effects on transcription factor binding, points to a causative model involving the gain of a BATF and JUND transcription binding site downstream of *CXCL12* capable of physically interacting with the *CXCL12* promoter. These results suggest an important modulating role of CXCL12 in the development of HIV-related NHL.

## Data Availability

Full summary statistics can be found in the GWAS catalog (https://www.ebi.ac.uk/gwas). The raw genotype data can be obtained through the respective cohorts.

## Acknowledgments

This study has been financed within the framework of the Swiss HIV Cohort Study, supported by the Swiss National Science Foundation (grant #177499), by SHCS project #789 and by the SHCS research foundation. The data are gathered by the Five Swiss University Hospitals, two Cantonal Hospitals, 15 affiliated hospitals and 36 private physicians (listed in http://www.shcs.ch/180-health-care-providers).

## Members of the Swiss HIV Cohort Study

Anagnostopoulos A, Battegay M, Bernasconi E, Böni J, Braun DL, Bucher HC, Calmy A, Cavassini M, Ciuffi A, Dollenmaier G, Egger M, Elzi L, Fehr J, Fellay J, Furrer H, Fux CA, Günthard HF (President of the SHCS), Haerry D (deputy of “Positive Council”), Hasse B, Hirsch HH, Hoffmann M, Hösli I, Huber M, Kahlert CR (Chairman of the Mother & Child Substudy), Kaiser L, Keiser O, Klimkait T, Kouyos RD, Kovari H, Ledergerber B, Martinetti G, Martinez de Tejada B, Marzolini C, Metzner KJ, Müller N, Nicca D, Paioni P, Pantaleo G, Perreau M, Rauch A (Chairman of the Scientific Board), Rudin C, Scherrer AU (Head of Data Centre), Schmid P, Speck R, Stöckle M (Chairman of the Clinical and Laboratory Committee), Tarr P, Trkola A, Vernazza P, Wandeler G, Weber R, Yerly S.

This work further benefited from the ANRS funding of both the Primo and Lymphovir cohorts.

Foundation Monahan and Fulbright funded the stay of CB at the National Cancer Institute (NCI).

The Genome-Wide Association Study (GWAS) of Non-Hodgkin Lymphoma (NHL) project was supported by the intramural program of the Division of Cancer Epidemiology and Genetics (DCEG), National Cancer Institute (NCI), National Institutes of Health (NIH). The datasets have been accessed through the NIH database for Genotypes and Phenotypes (dbGaP) under accession # phs000801. A full list of acknowledgements can be found in the supplementary note (Berndt SI et al., Nature Genet., 2013, PMID: 23770605).

## Authorship

C.W.T., J.F., P.J.M., C.S.R., C.B., C.H. and T.O.M. contributed to the conception and design of the study. C.W.T., J.F., P.J.M., F.A.S., D.C., L.M., C.G., I.T., S.K.H., M.C., A.R., M.B., M.H., P.S., E.B., H.F.G., C.S.R. and C.B. contributed to the acquisition of data. C.W.T., T.O.M., C.H., F.A.S., C.B., C.S.R. and J.F. contributed to the analysis and interpretation of data. C.W.T., J.F., C.S.R., C.B. and S.W. contributed to the drafting the article and revising it critically for important intellectual content.

All authors critically reviewed and approved the final manuscript.

Conflict of Interest Disclosure: Christian Hammer is a full-time employee of F. Hoffmann– La Roche/Genentech. The remaining authors declare no competing financial interests.

Correspondence: Jacques Fellay, School of Life Sciences, École Polytechnique Fédérale de Lausanne, Lausanne, Switzerland.

## References

1. Patel P, Hanson DL, Sullivan PS, et al. Incidence of Types of Cancer among HIV-Infected Persons Compared with the General Population in the United States, 1992–2003. Ann Intern Med. 2008;148(10):728–736.

2. Vogel M, Friedrich O, Lüchters G, et al. Cancer risk in HIV-infected individuals on HAART is largely attributed to oncogenic infections and state of immunocompetence. European Journal of Medical Research. 2011;16(3):101.

3. Robbins HA, Pfeiffer RM, Shiels MS, et al. Excess Cancers Among HIV-Infected People in the United States. J. Natl. Cancer Inst. 2015;107(4):.

4. Clifford GM, Polesel J, Rickenbach M, et al. Cancer Risk in the Swiss HIV Cohort Study: Associations With Immunodeficiency, Smoking, and Highly Active Antiretroviral Therapy. JNCI Journal of the National Cancer Institute. 2005;97(6):425–432.

5. Engels EA. Non-AIDS-defining malignancies in HIV-infected persons: etiologic puzzles, epidemiologic perils, prevention opportunities. AIDS. 2009;23(8):875–885.

6. Borges ÁH, Dubrow R, Silverberg MJ. Factors contributing to risk for cancer among HIV-infected individuals, and evidence that earlier combination antiretroviral therapy will alter this risk: Current Opinion in HIV and AIDS. 2014;9(1):34–40.

7. Guiguet M, Boué F, Cadranel J, et al. Effect of immunodeficiency, HIV viral load, and antiretroviral therapy on the risk of individual malignancies (FHDH-ANRS CO4): a prospective cohort study. The Lancet Oncology. 2009;10(12):1152–1159.

8. Shepherd L, Ryom L, Law M, et al. Differences in Virological and Immunological Risk Factors for Non-Hodgkin and Hodgkin Lymphoma. J Natl Cancer Inst. 2018;110(6):598–607.

9. Hleyhel M, Belot A, Bouvier AM, et al. Risk of AIDS-Defining Cancers Among HIV-1–Infected Patients in France Between 1992 and 2009: Results From the FHDH-ANRS CO4 Cohort. Clinical Infectious Diseases. 2013;57(11):1638–1647.

10. Robbins HA, Pfeiffer RM, Shiels MS, et al. Excess Cancers Among HIV-Infected People in the United States. JNCI: Journal of the National Cancer Institute. 2015;107(4):.

11. Swerdlow SH. WHO classification of tumours of haematopoietic and lymphoid tissues. International Agency for Research on Cancer; 2017.

12. McLaren PJ, Carrington M. The impact of host genetic variation on infection with HIV-1. Nat Immunol. 2015;16(6):577–583.

13. Sud A, Kinnersley B, Houlston RS. Genome-wide association studies of cancer: current insights and future perspectives. Nature Reviews Cancer. 2017;17(11):692–704.

14. Cerhan JR, Berndt SI, Vijai J, et al. Genome-wide association study identifies multiple susceptibility loci for diffuse large B cell lymphoma. Nature Genetics. 2014;46(11):1233–1238.

15. Conde L, Halperin E, Akers NK, et al. Genome-wide association study of follicular lymphoma identifies a risk locus at 6p21.32. Nat Genet. 2010;42(8):661–664.

16. Frampton M, da Silva Filho MI, Broderick P, et al. Variation at 3p24.1 and 6q23.3 influences the risk of Hodgkin’s lymphoma. Nature Communications. 2013;4:.

17. Kumar V, Matsuo K, Takahashi A, et al. Common variants on 14q32 and 13q12 are associated with DLBCL susceptibility. J Hum Genet. 2011;56(6):436–439.

18. Moutsianas L, Enciso-Mora V, Ma YP, et al. Multiple Hodgkin lymphoma– associated loci within the HLA region at chromosome 6p21.3. Blood. 2011;118(3):670–674.

19. Skibola CF, Bracci PM, Halperin E, et al. Genetic variants at 6p21.33 are associated with susceptibility to follicular lymphoma. Nat Genet. 2009;41(8):873–875.

20. Urayama KY, Jarrett RF, Hjalgrim H, et al. Genome-Wide Association Study of Classical Hodgkin Lymphoma and Epstein–Barr Virus Status–Defined Subgroups. JNCI J Natl Cancer Inst. 2012;104(3):240–253.

21. Vijai J, Kirchhoff T, Schrader KA, et al. Susceptibility Loci Associated with Specific and Shared Subtypes of Lymphoid Malignancies. PLoS Genetics. 2013;9(1):e1003220.

22. Skibola CF, Berndt SI, Vijai J, et al. Genome-wide Association Study Identifies Five Susceptibility Loci for Follicular Lymphoma outside the HLA Region. The American Journal of Human Genetics. 2014;95(4):462–471.

23. Vijai J, Wang Z, Berndt SI, et al. A genome-wide association study of marginal zone lymphoma shows association to the HLA region. Nat Commun. 2015;6:.

24. Tan DEK, Foo JN, Bei J-X, et al. Genome-wide association study of B cell non-Hodgkin lymphoma identifies 3q27 as a susceptibility locus in the Chinese population. Nature Genetics. 2013;45(7):804–807.

25. Schoeni-Affolter F, Ledergerber B, Rickenbach M, et al. Cohort Profile: The Swiss HIV Cohort Study. Int J Epidemiol. 2010;39(5):1179–1189.

26. Besson C, Lancar R, Prevot S, et al. Outcomes for HIV-associated diffuse large B-cell lymphoma in the modern combined antiretroviral therapy era. AIDS. 2017;31(18):2493.

27. Dalmasso C, Carpentier W, Meyer L, et al. Distinct Genetic Loci Control Plasma HIV-RNA and Cellular HIV-DNA Levels in HIV-1 Infection: The ANRS Genome Wide Association 01 Study. PLoS ONE. 2008;3(12):e3907.

28. Fellay J, Ge D, Shianna KV, et al. Common Genetic Variation and the Control of HIV-1 in Humans. PLOS Genetics. 2009;5(12):e1000791.

29. Loh P-R, Danecek P, Palamara PF, et al. Reference-based phasing using the Haplotype Reference Consortium panel. Nature Genetics. 2016;48(11):1443–1448.

30. Durbin R. Efficient haplotype matching and storage using the positional Burrows– Wheeler transform (PBWT). Bioinformatics. 2014;30(9):1266–1272.

31. McCarthy S, Das S, Kretzschmar W, et al. A reference panel of 64,976 haplotypes for genotype imputation. Nat Genet. 2016;advance online publication:

32. Chang CC, Chow CC, Tellier LC, et al. Second-generation PLINK: rising to the challenge of larger and richer datasets. Gigascience. 2015;4(1):1–16.

33. Price AL, Patterson NJ, Plenge RM, et al. Principal components analysis corrects for stratification in genome-wide association studies. Nat. Genet. 2006;38(8):904–909.

34. The International HapMap 3 Consortium. Integrating common and rare genetic variation in diverse human populations. Nature. 2010;467(7311):52–58.

35. Manichaikul A, Mychaleckyj JC, Rich SS, et al. Robust relationship inference in genome-wide association studies. Bioinformatics. 2010;26(22):2867–2873.

36. Yang J, Lee SH, Goddard ME, Visscher PM. GCTA: A Tool for Genome-wide Complex Trait Analysis. The American Journal of Human Genetics. 2011;88(1):76–82.

37. Yang J, Zaitlen NA, Goddard ME, Visscher PM, Price AL. Advantages and pitfalls in the application of mixed-model association methods. Nat Genet. 2014;46(2):100–106.

38. Kichaev G, Roytman M, Johnson R, et al. Improved methods for multi-trait fine mapping of pleiotropic risk loci. Bioinformatics. 2017;33(2):248–255.

39. Zhou J, Troyanskaya OG. Predicting effects of noncoding variants with deep learning–based sequence model. Nature Methods. 2015;12(10):931–934.

40. Khan A, Fornes O, Stigliani A, et al. JASPAR 2018: update of the open-access database of transcription factor binding profiles and its web framework. Nucleic Acids Res. 2018;46(D1):D260–D266.

41. Rao SSP, Huntley MH, Durand NC, et al. A 3D Map of the Human Genome at Kilobase Resolution Reveals Principles of Chromatin Looping. Cell. 2014;159(7):1665–1680.

42. Wang Y, Song F, Zhang B, et al. The 3D Genome Browser: a web-based browser for visualizing 3D genome organization and long-range chromatin interactions. Genome Biology. 2018;19(1):151.

43. Schofield EC, Carver T, Achuthan P, et al. CHiCP: a web-based tool for the integrative and interactive visualization of promoter capture Hi-C datasets. Bioinformatics. 2016;32(16):2511–2513.

44. Mifsud B, Tavares-Cadete F, Young AN, et al. Mapping long-range promoter contacts in human cells with high-resolution capture Hi-C. Nature Genetics. 2015;47(6):598–606.

45. Cairns J, Freire-Pritchett P, Wingett SW, et al. CHiCAGO: robust detection of DNA looping interactions in Capture Hi-C data. Genome Biol. 2016;17(1):127.

46. Lappalainen T, Sammeth M, Friedländer MR, et al. Transcriptome and genome sequencing uncovers functional variation in humans. Nature. 2013;501(7468):506–511.

47. Piasecka B, Duffy D, Urrutia A, et al. Distinctive roles of age, sex, and genetics in shaping transcriptional variation of human immune responses to microbial challenges. PNAS. 2018;115(3):E488–E497.

48. GTEx Consortium. Genetic effects on gene expression across human tissues. Nature. 2017;550(7675):204–213.

49. Alpern D, Gardeux V, Russeil J, et al. BRB-seq: ultra-affordable high-throughput transcriptomics enabled by bulk RNA barcoding and sequencing. Genome Biology. 2019;20(1):71.

50. Berndt SI, Skibola CF, Joseph V, et al. Genome-wide association study identifies multiple risk loci for chronic lymphocytic leukemia. Nature Genetics. 2013;45(8):868–876.

51. Johnson JL, Abecasis GR. GAS Power Calculator: web-based power calculator for genetic association studies. bioRxiv. 2017;164343.

52. Mohammadi P, Desfarges S, Bartha I, et al. 24 Hours in the Life of HIV-1 in a T Cell Line. PLOS Pathogens. 2013;9(1):e1003161.

53. Lim U, Kocarnik JM, Bush WS, et al. Pleiotropy of Cancer Susceptibility Variants on the Risk of Non-Hodgkin Lymphoma: The PAGE Consortium. PLoS ONE. 2014;9(3):e89791.

54. Sei S, O’Neill DP, Stewart SK, et al. Increased Level of Stromal Cell-Derived Factor-1 mRNA in Peripheral Blood Mononuclear Cells from Children with AIDS-related Lymphoma. Cancer Res. 2001;61(13):5028–5037.

55. Rabkin CS, Yang Q, Goedert JJ, et al. Chemokine and Chemokine Receptor Gene Variants and Risk of Non-Hodgkin’s Lymphoma in Human Immunodeficiency Virus-1–Infected Individuals. Blood. 1999;93(6):1838–1842.

56. Meng W, Xue S, Chen Y. The role of CXCL12 in tumor microenvironment. Gene. 2018;641:105–110.

57. Peled A, Klein S, Beider K, Burger JA, Abraham M. Role of CXCL12 and CXCR4 in the pathogenesis of hematological malignancies. Cytokine. 2018;109:11–16.

58. Piovan E, Tosello V, Indraccolo S, et al. Chemokine receptor expression in EBV-associated lymphoproliferation in hu/SCID mice: implications for CXCL12/CXCR4 axis in lymphoma generation. Blood. 2005;105(3):931–939.

59. Strober BJ, Elorbany R, Rhodes K, et al. Dynamic genetic regulation of gene expression during cellular differentiation. Science. 2019;364(6447):1287–1290.

60. Quigley M, Pereyra F, Nilsson B, et al. Transcriptional analysis of HIV-specific CD8+ T cells shows that PD-1 inhibits T cell function by upregulating BATF. Nat. Med. 2010;16(10):1147–1151.

61. Gasser O, Bihl FK, Wolbers M, et al. HIV Patients Developing Primary CNS Lymphoma Lack EBV-Specific CD4þ T Cell Function Irrespective of Absolute CD4þ T Cell Counts. PLoS Medicine. 2007;4(3):6.

62. SH S, E C, NL H, et al. WHO Classification of Tumours of Haematopoietic and Lymphoid Tissues.

